# Examining Health Service Rates Among Residents of Retirement Homes and Other Older Adult Populations in Ontario, Canada: A Population-Based Cohort Study

**DOI:** 10.1101/2021.09.17.21263742

**Authors:** Derek R. Manis, Jeffrey W. Poss, Aaron Jones, Paula A. Rochon, Susan E. Bronskill, Michael A. Campitelli, Richard Perez, Nathan M. Stall, Ahmad Rahim, Glenda Babe, Jean-Éric Tarride, Julia Abelson, Andrew P. Costa

## Abstract

**Background:** There are no standardized reporting systems or assessments specific to residents of retirement homes in North America. As such, little is known about these older adults as a distinct population. We created a new population-level cohort of residents of retirement homes and examined their health service rates relative to other older adult populations.

**Methods:** We conducted a population-based retrospective cohort study in Ontario, Canada in 2018. The postal codes of all licensed retirement homes (*n* = 757) were classified and linked to individual-level health system administrative data to derive a cohort of residents of retirement homes. A generalized linear model with a gamma distribution and log link function was used to model rates of emergency department visits, hospitalizations, alternate levels of care (ALC) days, primary care visits, and specialist physician visits.

**Results:** Residents of retirement homes comprised two percent of the older adult population in Ontario (*n* = 54,773; 2.3%). After adjustment for relevant characteristics, residents of retirement homes had 10 times the rate of emergency department visits (Relative Rate [RR] 10.02, 95% Confidence Interval [CI] 9.83 to 10.21), 20 times the rate of hospitalizations (RR 20.43, 95% CI 20.08 to 20.78), and 44 times the rate ALC days (RR 43.91, 95% CI 43.28 to 44.54) compared to community-dwelling older adults.

**Interpretation:** Residents of retirement homes are a distinct older adult population with high rates of hospital-based care. Our findings contribute to policy debates about the provision of health care in privately operated congregate care settings for older adults.

## INTRODUCTION

Retirement homes are private, congregate living environments that deliver supportive care to adults who are 65 years of age and older (1,2). Retirement homes are often marketed to provide a lifestyle and community, and these homes provide a range of assisted living care services (e.g., meals, administration of medication, nursing services, etc.) on a cost recovery basis (2). These homes predominately operate on a private, for-profit business model, and the room, board, and services are purchased by residents and/or their family or friend caregivers (1,2).

Retirement homes are referred to as assisted living facilities in other North American jurisdictions. The legislative and regulatory operating requirements for these homes vary (3); Ontario is the only jurisdiction to regulate and license retirement homes through an independent, not-for-profit regulator (i.e., Retirement Homes Regulatory Authority [RHRA]) (2). There are more than 700 licensed retirement homes in Ontario that can house over 70,000 older adults, which is comparable to the number of beds in the long-term care home sector (1,2,4). Unlike long-term care homes where the validated Resident Assessment Instrument Minimum Data Set (RAI-MDS) is used, there are no standardized assessments specific to residents of retirement homes (5). The absence of standardized assessments that are periodically conducted on residents of retirement homes, coupled with the privately financed nature of the sector, presents unique challenges to identify these older adults using health system administrative data and understand their health service use relative to other older adult populations.

While there is a body of research describing the assisted living sector and residents of assisted living facilities in the United States (6–12), the Canadian literature requires a more in-depth investigation. Canadian studies have investigated transitions to a long-term care home, risk of hospitalization among those who live with dementia, and events and health conditions associated with the transition to a congregate care setting for older adults (13–17). To date, no population- or provincial-level cohort of residents of retirement homes in Canada has been created to define the retirement home and assisted living sector, describe the characteristics and health service use of older adults who purchase these services, and position the sector in the gradient of care services and housing needs for older adults. In this study, we create a new population-level cohort of residents of retirement homes and examine their health service rates relative to other older adult populations (i.e., residents of long-term care homes, home care recipients in the community, and community-dwelling older adults) in Ontario, Canada.

## METHODS

### Study Design and Setting

We conducted a population-based retrospective cohort study using linked, individual-level health system administrative data in 2018 in Ontario, Canada at ICES. ICES is an independent, non-profit research institute whose legal status under Ontario’s health information privacy law allows it to collect and analyze health care and demographic data, without consent, for health system evaluation and improvement. The use of the data in this project is authorized under section 45 of Ontario’s *Personal Health Information Protection Act* (PHIPA) and does not require review by a Research Ethics Board. We followed the REporting of studies Conducted using Observational Routinely-collected health Data (RECORD) statement guideline (Supplemental Table 1) (18).

### Data

The RHRA shared their public register of licensed retirement homes, which contains historical data on the license of the home, resident and suite capacities, provision of regulated care services, and full postal address. There were 757 licensed retirement homes in 2018 (Supplemental Table 2). We verified and visualized the postal code of each licensed retirement home through a variety of sources (i.e., Canada Post, Statistics Canada, and Google Maps). Building off research that examined the feasibility of using postal codes to identify residents of retirement homes (19), we used a modified taxonomy to classify the postal code of each licensed retirement home as unique, or not unique, to the retirement home. We imported the RHRA’s public register and our classified postal code data on licensed retirement homes to ICES. The health system administrative datasets used are listed and described in Supplemental Table 3. These datasets were linked using unique encoded identifiers and analyzed at ICES.

### Identification of Residents of Retirement Homes

We identified adults who were 65 years of age of and older and had a postal code that ever matched to a licensed retirement home with a unique postal code classification in 2018 (Figure 1). There were substantially more adults who were 65 years of age and older and had a postal code that matched to a licensed retirement home with a not unique postal code classification than beds in licensed retirement homes. We limited our identification of residents of retirement homes with not unique postal codes to those who received home care services in a retirement home, as the receipt of home care services specified whether the services were provided in a retirement home. We defined the index date as when the adults’ postal code matched to the postal code of a licensed retirement home; we terminated follow-up when the adult transitioned to a long-term care home, complex continuing care facility, or died.

**Figure 1.**
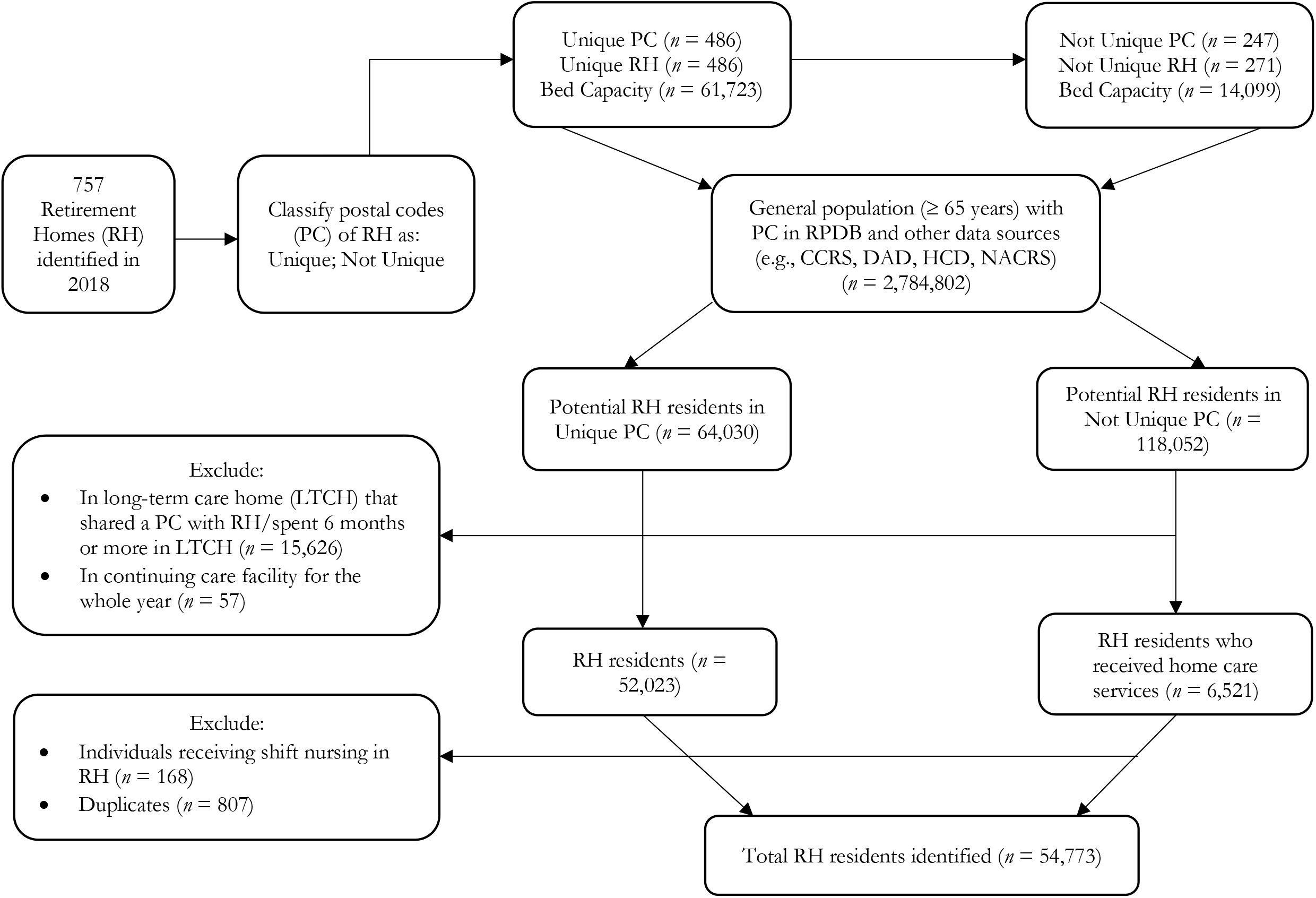
Creation of the Residents of Retirement Homes Cohort in 2018 (*n* = 54,773)

We excluded adults who were institutionalized in a long-term care home that was co-located with a retirement home and adults who resided in a long-term care home for more than half of 2018 (i.e., six months plus one day) (*n* = 15,626). Adults who resided in a continuing care facility for the whole year in 2018 were excluded (*n* = 57). We also excluded adults who received shift nursing in retirement homes (*n* = 168), as these individuals were temporarily housed in the retirement home through convalescent government programs, and so these individuals were not true residents of retirement homes. We excluded duplicate adults who moved from one retirement home to another during the year (*n =* 807). According to the RHRA’s register, there were 75,822 beds in all licensed retirement homes in 2018, and our approach identified a cohort of 54,733 residents of retirement homes (*n =* 54,733; 72.2%).

### Identification of Other Older Adult Populations

For comparison purposes, we identified residents of long-term care homes, older adults who received home care services in the community, and community-dwelling adults who were 65 years of age and older in 2018. Residents of long-term care homes were identified by their inclusion in the Continuing Care Reporting System (*n* = 96,528). Adults who received home care services in their community were differentiated from residents of retirement homes by their postal code that never matched to a postal code associated with a licensed retirement home (*n* = 290,245). Community-dwelling adults were defined as those who were 65 years of age and older and never met any of the above criteria (*n* = 1,967,612). We defined the index date as when the adult met the criteria to be categorized as one of the mutually exclusive older adult populations in 2018; we terminated follow-up when the adult met the criteria to be categorized as a different older adult population or died.

### Measures

The outcomes of interest were annual rates of emergency department visits, hospitalizations, alternate levels of care (ALC) days, primary care visits, and specialist physician visits. These rates were standardized at the level of the individual (i.e., from index to end of follow-up). Emergency department visits were defined as any care received in an emergency department. Hospitalizations were defined as any hospitalization. ALC days were obtained from the Discharge Abstract Database.

Primary care visits among residents of retirement homes, home care recipients in the community, and community-dwelling older adults were defined as any billing by a family or community medicine physician to the universal health insurance plan where the location of the visit occurred in an office, home, or via the telephone. Primary care visits among residents of long-term care homes were similarly defined in accordance with the Monthly Management System and included a long-term care home as the visit location. Specialist physician visits were defined as any physician billing whose speciality was not family or community medicine to the universal health insurance plan. All individuals could only have one primary care and/or specialist physician visit per physician per day.

Sociodemographic (i.e., age and sex) and community characteristics (i.e., urban location, neighborhood income quintile, Ontario Marginalization Index) were obtained at the index date. Clinical comorbidities were also obtained at the index date from physician-diagnosed billing codes to the universal health insurance plan in Ontario, ICD-9 or ICD-10 diagnosis codes, and validated ICES-derived cohorts (20–27).

### Statistical Analysis

Counts and proportions were calculated for categorical sociodemographic, community, and clinical variables; means and standard deviations were calculated for continuous, normally distributed sociodemographic variables, and medians and interquartile ranges were calculated for continuous, not normally distributed sociodemographic variables. A generalized linear model with a gamma distribution and log link function was used to model the standardized health service rates among the different older adult populations, and community-dwelling older adults were used as the reference population. The gamma distribution and log link function is appropriate to model dispersed rates and/or costs in dollars (28). Crude and adjusted relative rates and 95% confidence intervals were calculated from exponentiated beta coefficients. All statistical tests were two-tailed, and the level of statistical significance was *P* < .05. Variance inflation factors were calculated to assess for multicollinearity. A sex-stratified subgroup analysis was conducted. Dataset processing and statistical analyses were conducted in SAS Enterprise 9.4 (Cary, NC, USA).

## RESULTS

Residents of retirement homes comprised two percent of adults aged 65 and older in Ontario in 2018 (*n* = 54,773; 2.3%). More than two thirds of residents of retirement homes were female (*n* = 37,768; 69.0%), and residents of retirement homes had a mean age of 87.7 years (Table 1). Hypertension (*n* = 47,212; 86.2%), osteoarthritis (*n* = 36,978; 67.5%), mood disorders (*n* = 35,000; 63.9%) and dementia (*n* = 20,651; 37.7%) were the most prevalent clinical comorbidities among residents of retirement homes. More than 90% of all residents of retirement homes resided in urban communities (*n =* 50,650; 92.5%).

**Table 1.** Sociodemographic Characteristics and Clinical Comorbidities Among Residents of Retirement Homes, Residents of Long-Term Care Homes, Home Care Recipients in the Community, and Community-Dwelling Older Adults in 2018 (*n* = 2,419,158)

|  | **Residents of Retirement Homes** | **Residents of Long-Term Care Homes** | **Home Care Recipients in the Community** | **Community-Dwelling Older Adults** |
| --- | --- | --- | --- | --- |
| *n* (%) | 54,773 (2.3) | 96,528 (4.0) | 290,245 (12.0) | 1,967,612 (81.6) |
| **Demographic Characteristics, *n* (%)** |  |  |  |  |
| Age, mean (SD) | 87.7 (7.21) | 85.3 (8.22) | 79.3 (8.41) | 73.8 (6.68) |
| Female | 37,768 (69.0) | 66,097 (68.5) | 163,216 (56.2) | 1,046,805 (53.2) |
| **Clinical Comorbidities, *n* (%)** |  |  |  |  |
| Asthma | 8,334 (15.2) | 13,903 (14.4) | 52,169 (18.0) | 250,749 (12.7) |
| Cancer | 14,557 (26.6) | 19,578 (20.3) | 93,550 (32.2) | 316,863 (16.1) |
| Cardiac Arrhythmias | 19,723 (36.0) | 28,585 (29.6) | 90,111 (31.0) | 320,234 (16.3) |
| Cerebrovascular Accident | 11,796 (21.5) | 26,781 (27.7) | 51,683 (17.8) | 109,713 (5.6) |
| Chronic Coronary Disease | 23,210 (42.4) | 39,091 (40.5) | 119,603 (41.2) | 494,772 (25.1) |
| Chronic Obstructive Pulmonary Disease | 17,486 (31.9) | 31,033 (32.1) | 97,929 (33.7) | 370,765 (18.8) |
| Congestive Heart Failure | 16,063 (29.3) | 25,119 (26.0) | 73,498 (25.3) | 130,132 (6.6) |
| Dementia | 20,651 (37.7) | 76,485 (79.2) | 52,952 (18.2) | 51,372 (2.6) |
| Diabetes | 17,097 (31.2) | 36,513 (37.8) | 119,516 (41.2) | 566,716 (28.8) |
| Hypertension | 47,212 (86.2) | 80,882 (83.8) | 240,470 (82.9) | 1,317,840 (67.0) |
| Mood Disorders | 35,000 (63.9) | 66,365 (68.8) | 168,785 (58.2) | 951,562 (48.4) |
| Myocardial Infarction | 6,542 (11.9) | 10,554 (10.9) | 35,141 (12.1) | 116,275 (5.9) |
| Osteoarthritis | 36,978 (67.5) | 65,335 (67.7) | 169,423 (58.4) | 794,611 (40.4) |
| Osteoporosis | 8,334 (15.2) | 13,903 (14.4) | 49,938 (16.9) | 261,113 (13.3) |
| Other Mental Health | 16,097 (29.4) | 34,350 (35.6) | 94,282 (32.5) | 475,043 (24.1) |
| Renal Disease | 8,236 (15.0) | 13,487 (14.0) | 50,374 (17.4) | 118,137 (6.0) |
| Rheumatoid Arthritis | 2,091 (3.8) | 3,261 (3.4) | 12,639 (4.4) | 46,713 (2.4) |
| **Community Characteristics, *n* (%)** |  |  |  |  |
| Urban Location | 50,650 (92.5) | 83,171 (86.2) | 253,336 (87.3) | 1,712,634 (87.0) |
| **Neighborhood Income Quintile** |  |  |  |  |
| 1 (Least) | 13,037 (23.8) | 28,683 (29.7) | 72,124 (24.9) | 370,961 (18.9) |
| 2 | 12,685 (23.2) | 21,047 (21.8) | 64,141 (22.1) | 405,261 (20.6) |
| 3 | 10,405 (19.0) | 16,892 (17.5) | 56,242 (19.4) | 396,048 (20.1) |
| 4 | 10,409 (19.0) | 15,936 (16.5) | 48,907 (16.9) | 378,654 (19.2) |

|  | **Residents of Retirement Homes** | **Residents of Long-Term Care Homes** | **Home Care Recipients in the Community** | **Community-Dwelling Older Adults** |
| --- | --- | --- | --- | --- |
| 5 (Highest) | 7,639 (13.9) | 13,291 (13.8) | 48,093 (16.6) | 412,074 (20.9) |
| Missing | 598 (1.1) | 679 (0.7) | 728 (0.3) | 4,614 (0.2) |
| Ontario Marginalization Index Summary Score, median (IQR) | 3 (3 to 4) | 4 (3 to 4) | 3 (3 to 4) | 3 (3 to 4) |

Abbreviations: IQR, Interquartile Range; SD, Standard Deviation

The crude and sex-stratified health service rates are described in Supplemental Table 4. After adjustment for sociodemographic characteristics and clinical comorbidities at index, residents of retirement homes had 10 times the rate of emergency department visits (Relative Rate [RR] 10.02, 95% Confidence Interval [CI], 9.83 to 10.21), 20 times the rate of hospitalizations (RR 20.43, 95% CI 20.08 to 20.78), 44 times the rate of ALC days (RR 43.91, 95% CI 43.28 to 44.54), and nearly twice the rate of primary care (RR 1.99, 95% CI 1.97 to 2.02) and specialist physician visits (RR 1.62, 95% CI 1.59 to 1.65), compared to community-dwelling older adults (Table 2). Male residents of retirement homes had higher rates of emergency department visits, hospitalizations, and ALC days than female residents of retirement homes, but similar rates of primary care and specialist physician visits (Table 3). Adjusted beta coefficients and standard errors are available in Supplementary Tables 5 and 6.

**Table 2.**
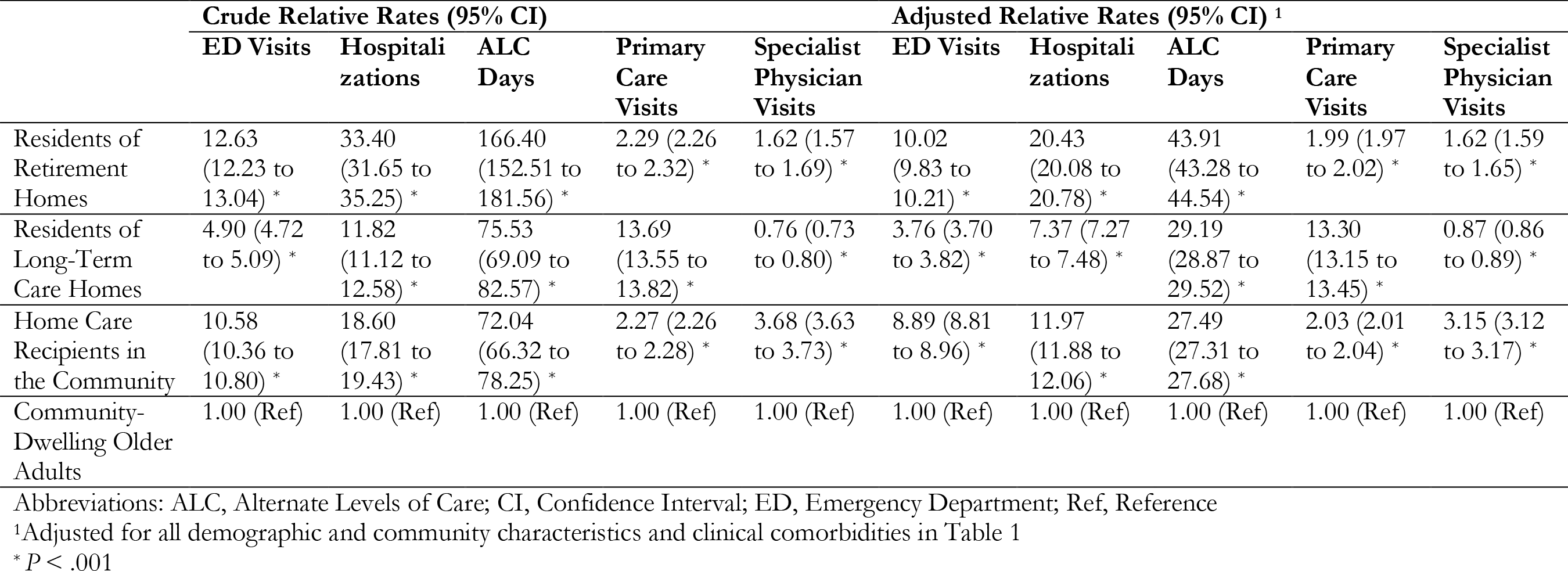
Annual, Standardized Health Service Rates Among Residents of Retirement Homes, Residents of Long-Term Care Homes, and Home Care Recipients in the Community in 2018

|  | Crude Relative Rates (95% CI) |  |  |  |  | Adjusted Relative Rates (95% CI) <sup>1</sup> |  |  |  |  |
| --- | --- | --- | --- | --- | --- | --- | --- | --- | --- | --- |
|  | ED Visits | Hospitalizations | ALC Days | Primary Care Visits | Specialist Physician Visits | ED Visits | Hospitalizations | ALC Days | Primary Care Visits | Specialist Physician Visits |
| Residents of Retirement Homes | 12.63<br>(12.23 to 13.04) * | 33.40<br>(31.65 to 35.25) * | 166.40<br>(152.51 to 181.56) * | 2.29 (2.26 to 2.32) * | 1.62 (1.57 to 1.69) * | 10.02<br>(9.83 to 10.21) * | 20.43<br>(20.08 to 20.78) * | 43.91<br>(43.28 to 44.54) * | 1.99 (1.97 to 2.02) * | 1.62 (1.59 to 1.65) * |
| Residents of Long-Term Care Homes | 4.90 (4.72 to 5.09) * | 11.82<br>(11.12 to 12.58) * | 75.53<br>(69.09 to 82.57) * | 13.69<br>(13.55 to 13.82) * | 0.76 (0.73 to 0.80) * | 3.76 (3.70 to 3.82) * | 7.37 (7.27 to 7.48) * | 29.19<br>(28.87 to 29.52) * | 13.30<br>(13.15 to 13.45) * | 0.87 (0.86 to 0.89) * |
| Home Care Recipients in the Community | 10.58<br>(10.36 to 10.80) * | 18.60<br>(17.81 to 19.43) * | 72.04<br>(66.32 to 78.25) * | 2.27 (2.26 to 2.28) * | 3.68 (3.63 to 3.73) * | 8.89 (8.81 to 8.96) * | 11.97<br>(11.88 to 12.06) * | 27.49<br>(27.31 to 27.68) * | 2.03 (2.01 to 2.04) * | 3.15 (3.12 to 3.17) * |
| Community-Dwelling Older Adults | 1.00 (Ref) | 1.00 (Ref) | 1.00 (Ref) | 1.00 (Ref) | 1.00 (Ref) | 1.00 (Ref) | 1.00 (Ref) | 1.00 (Ref) | 1.00 (Ref) | 1.00 (Ref) |

Abbreviations: ALC, Alternate Levels of Care; CI, Confidence Interval; ED, Emergency Department; Ref, Reference
<sup>1</sup>Adjusted for all demographic and community characteristics and clinical comorbidities in Table 1\* $P < .001$

**Table 3.** Sex-Stratified Subgroup Analysis of Annual, Standardized Health Service Rates Among Residents of Retirement Homes, Residents of Long-Term Care Homes, and Home Care Recipients in the Community in 2018

|  |  | Crude Relative Rates (95% CI) |  |  |  |  | Adjusted Relative Rates (95% CI) <sup>1</sup> |  |  |  |  |
| --- | --- | --- | --- | --- | --- | --- | --- | --- | --- | --- | --- |
|  |  | ED Visits | Hospitalizations | ALC Days | Primary Care Visits | Specialist Physician Visits | ED Visits | Hospitalizations | ALC Days | Primary Care Visits | Specialist Physician Visits |
| Males (*n* = 1,094,635) | Residents of Retirement Homes | 15.57<br>(15.08 to 16.07) * | 35.69<br>(34.63 to 36.78) * | 148.40<br>(144.46 to 152.44) * | 2.48<br>(2.42 to 2.54) * | 1.90<br>(1.84 to 1.95) * | 12.15<br>(11.76 to 12.56) * | 22.30<br>(21.63 to 22.99) * | 51.07<br>(49.81 to 52.37) * | 2.03<br>(1.98 to 2.07) * | 1.64<br>(1.60 to 1.69) * |
|  | Residents of Long-Term Care Homes | 6.32<br>(6.17 to 6.47) * | 12.97<br>(12.68 to 13.27) * | 83.60<br>(81.92 to 85.31) * | 14.67<br>(14.42 to 14.93) * | 0.87<br>(0.85 to 0.89) * | 4.88<br>(4.75 to 5.01) * | 8.77<br>(8.56 to 9.00) * | 58.72<br>(57.56 to 59.89) * | 13.61<br>(13.34 to 13.88) * | 0.86<br>(0.84 to 0.88) * |
|  | Home Care Recipients in the Community | 12.89<br>(12.73 to 13.04) * | 19.05<br>(18.83 to 19.27) * | 60.19<br>(59.57 to 60.82) * | 2.54<br>(2.51 to 2.56) * | 4.03<br>(3.99 to 4.08) * | 11.19<br>(11.06 to 11.33) * | 13.45<br>(13.29 to 13.60) * | 31.77<br>(31.46 to 32.09) * | 2.25<br>(2.23 to 2.27) * | 3.35<br>(3.32 to 3.39) * |
|  | Community-Dwelling Older Adults | 1.00<br>(Ref) | 1.00<br>(Ref) | 1.00<br>(Ref) | 1.00<br>(Ref) | 1.00<br>(Ref) | 1.00<br>(Ref) | 1.00<br>(Ref) | 1.00<br>(Ref) | 1.00<br>(Ref) | 1.00<br>(Ref) |
| Females (*n* = 1,313,038) | Residents of Retirement Homes | 11.11<br>(10.87 to 11.36) * | 30.92<br>(30.30 to 31.54) * | 120.23<br>(118.08 to 122.41) * | 2.18<br>(2.15 to 2.22) * | 1.52<br>(1.49 to 1.55) * | 8.89<br>(8.69 to 9.10) * | 19.38<br>(18.98 to 19.78) * | 40.09<br>(39.39 to 40.80) * | 2.00<br>(1.96 to 2.03) * | 1.66<br>(1.62 to 1.69) * |
|  | Residents of Long-Term Care Homes | 4.19<br>(4.12 to 4.26) * | 10.78<br>(10.62 to 10.95) * | 45.62<br>(45.00 to 46.26) * | 13.09<br>(12.93 to 13.24) * | 0.73<br>(0.72 to 0.74) * | 3.17<br>(3.11 to 3.24) * | 6.61<br>(6.50 to 6.73) * | 15.26<br>(15.05 to 15.48) * | 13.31<br>(13.12 to 13.50) * | 0.92<br>(0.90 to 0.93) * |
|  | Home Care Recipients in the Community | 8.61<br>(8.51 to 8.70) * | 16.19<br>(16.03 to 16.36) * | 50.41<br>(49.94 to 50.87) * | 2.07<br>(2.05 to 2.08) * | 3.40<br>(3.36 to 3.43) * | 7.17<br>(7.09 to 7.25) * | 10.82<br>(10.71 to 10.93) * | 24.02<br>(23.80 to 24.23) * | 1.86<br>(1.85 to 1.88) * | 3.01<br>(2.98 to 3.04) * |
|  | Community-Dwelling Older Adults | 1.00<br>(Ref) | 1.00<br>(Ref) | 1.00<br>(Ref) | 1.00<br>(Ref) | 1.00<br>(Ref) | 1.00<br>(Ref) | 1.00<br>(Ref) | 1.00<br>(Ref) | 1.00<br>(Ref) | 1.00<br>(Ref) |

Abbreviations: ALC, Alternate Levels of Care; CI, Confidence Interval; ED, Emergency Department; Ref, Reference
<sup>1</sup>Adjusted for all demographic and community characteristics and clinical comorbidities in Table 1
\* $P < .001$

## INTERPRETATION

Residents of retirement homes had the highest rates of emergency department visits, hospitalizations, and ALC days relative to the other older adult populations in Ontario, Canada in 2018. These older adults purchase health care services from their retirement home to support independent living, yet we found they consume more publicly funded hospital-based care and have lower rates of primary care and specialist physician visits. Our findings contribute to on-going policy debates about privately financed and delivered health care, and the provision of health care in privately operated congregate care settings for older adults, in jurisdictions that provide universal health insurance to its citizens (29,30).

The variation in legislative and operating requirements for retirement homes has been shown to affect rates of hospital-based use among these residents (31,32). Previous studies found residents of retirement homes have substantially higher rates of emergency department visits and hospitalizations compared to community-dwelling older adults and residents of long-term care homes (11,33). Our findings align with the literature and may suggest residents of retirement homes have higher needs for care compared to other older adult populations.

Residents of retirement homes had the highest rates of ALC days relative to the other older adult populations in our study, which suggests the needs of some residents may exceed the capacity of their home to provide the level and scope of care needed. Moreover, some of these residents may not be able to afford additional care from their retirement home, as rates for heavy care in Ontario can exceed $6,000 per month (34). Nearly half of the residents of retirement homes lived in middle- and low-income neighborhoods. The costs for heavy care are likely out of reach for many of these older adults (35–37), which underscores the need for equitable policies that support health and housing for older adults.

Residents of retirement homes had lower rates of primary care and specialist physician visits relative to home care recipients who lived in the community. Residents of long-term care homes receive primary care through the Monthly Management System, but no similar model exists for retirement homes. Our findings suggest the implementation and expansion of similar medical models of care in retirement homes may be an important intervention to promote continuity of care and reduce rates of hospital-based care among this population.

Retirement home and assisted living markets in North America are rapidly expanding to accommodate the varying preferences of older adults for housing, health, and social care (2,5– 7,35,38). The growth and availability of beds in retirement homes and assisted living facilities outpaces that of long-term care homes (2,38), and this growth is likely attributed to fewer legislative and regulatory requirements than long-term care homes. The increased supply of retirement homes may suggest that retirement homes are a substitute for a long-term care home (7,38), which suggests that retirement homes are an important link in the continuum of care settings for older adults and should be subject to similar regulatory oversight.

There are limitations to our study. We conducted a secondary analysis of health system administrative data; as such, there is the possibility of misclassification bias and residual confounding could influence our results and interpretation. We were unable to identify residents of retirement homes who did not receive home care services in licensed retirement homes with not unique postal codes. We were also unable to determine the occupancy of each retirement home, as the RHRA does not require operators to disclose this statistic as a condition for licensing (2). As the occupancy of each retirement home is unknown, the size of our cohort of residents of retirement homes may reflect the actual population size.

Residents of retirement home are a unique older adult population in Ontario, Canada. Future research should examine with more granularity the reasons why residents of retirement homes visited emergency departments and/or were hospitalized to understand their needs for hospital-based care that may not be met in their retirement home.

## Data Availability

The dataset from this study is held securely in coded form at ICES. While data sharing agreements prohibit ICES from making the data set publicly available, access may be granted to those who meet pre-specified criteria for confidential access, available at www.ices.on.ca/DAS. The full data set creation plan and underlying analytic code are available from the corresponding author upon request, understanding that the programs may rely upon coding templates or macros that are unique to ICES.

## ACKNOWLEDGMENTS

This study was supported by ICES, which is funded by an annual grant from the Ontario Ministry of Health (MOH) and the Ministry of Long-Term Care (MLTC). Parts of this material are based on data and/or information compiled and provided by CIHI. The analyses, conclusions, opinions, and statements expressed herein are solely those of the authors and do not reflect those of the funding or data sources; no endorsement is intended or should be inferred. We extend our gratitude to Paul Pham, Adriane Castellino, and Chloe Ma at the Retirement Homes Regulatory Authority for sharing the registry of licensed retirement homes.

## SUPPLEMENTARY MATERIAL

**Supplemental Table 1.** RECORD Checklist

|  | Item No. | STROBE items | Location in manuscript where items are reported | RECORD items | Location in manuscript where items are reported |
| --- | --- | --- | --- | --- | --- |
| **Title and Abstract** |  |  |  |  |  |
|  | 1 | (a) Indicate the study's design with a commonly used term in the title or the abstract (b) Provide in the abstract an informative and balanced summary of what was done and what was found | p. 1,2 | RECORD 1.1: The type of data used should be specified in the title or abstract. When possible, the name of the databases used should be included.<br>RECORD 1.2: If applicable, the geographic region and timeframe within which the study took place should be reported in the title or abstract.<br>RECORD 1.3: If linkage between databases was conducted for the study, this should be clearly stated in the title or abstract. | p. 1,2 |
| **Introduction** |  |  |  |  |  |
| Background rationale | 2 | Explain the scientific background and rationale for the investigation being reported | p. 3,4 |  |  |
| Objectives | 3 | State specific objectives, including any prespecified hypotheses | p. 4 |  |  |
| **Methods** |  |  |  |  |  |
| Study Design | 4 | Present key elements of study design early in the paper | p. 4 |  |  |

**Supplemental Table 1.** RECORD Checklist
|  | Item No. | STROBE items | Location in manuscript where items are reported | RECORD items | Location in manuscript where items are reported |
| --- | --- | --- | --- | --- | --- |
| Setting | 5 | Describe the setting, locations, and relevant dates, including periods of recruitment, exposure, follow-up, and data collection | p. 4 |  |  |
| Participants | 6 | *(a) Cohort study* - Give the eligibility criteria, and the sources and methods of selection of participants. Describe methods of follow-up<br>*Case-control study* - Give the eligibility criteria, and the sources and methods of case ascertainment and control selection. Give the rationale for the choice of cases and controls<br>*Cross-sectional study* - Give the eligibility criteria, and the sources and methods of selection of participants<br>*(b) Cohort study* - For matched studies, give matching criteria and number of exposed and unexposed | p. 5,6 | RECORD 6.1: The methods of study population selection (such as codes or algorithms used to identify subjects) should be listed in detail. If this is not possible, an explanation should be provided.<br>RECORD 6.2: Any validation studies of the codes or algorithms used to select the population should be referenced. If validation was conducted for this study and not published elsewhere, detailed methods and results should be provided.<br>RECORD 6.3: If the study involved linkage of databases, consider use of a flow diagram or other graphical display to demonstrate the data linkage process, including the number of individuals with linked data at each stage. | p. 5,6,16 |

**Supplemental Table 1.** RECORD Checklist
|  | Item No. | STROBE items | Location in manuscript where items are reported | RECORD items | Location in manuscript where items are reported |
| --- | --- | --- | --- | --- | --- |
|  |  | *Case-control study* - For matched studies, give matching criteria and the number of controls per case |  |  |  |
| Variables | 7 | Clearly define all outcomes, exposures, predictors, potential confounders, and effect modifiers. Give diagnostic criteria, if applicable. | p. 6,7 | RECORD 7.1: A complete list of codes and algorithms used to classify exposures, outcomes, confounders, and effect modifiers should be provided. If these cannot be reported, an explanation should be provided. | p. 6,7 |
| Data sources/<br>measurement | 8 | For each variable of interest, give sources of data and details of methods of assessment (measurement). Describe comparability of assessment methods if there is more than one group | p. 6,7 |  |  |
| Bias | 9 | Describe any efforts to address potential sources of bias |  |  |  |
| Study size | 10 | Explain how the study size was arrived at |  |  |  |
| Quantitative variables | 11 | Explain how quantitative variables were handled in the analyses. If applicable, | p. 6,7 |  |  |

**Supplemental Table 1.** RECORD Checklist
|  | Item No. | STROBE items | Location in manuscript where items are reported | RECORD items | Location in manuscript where items are reported |
| --- | --- | --- | --- | --- | --- |
|  |  | describe which groupings were chosen, and why |  |  |  |
| Statistical methods | 12 | (a) Describe all statistical methods, including those used to control for confounding<br>(b) Describe any methods used to examine subgroups and interactions<br>(c) Explain how missing data were addressed<br>(d) *Cohort study* - If applicable, explain how loss to follow-up was addressed<br>*Case-control study* - If applicable, explain how matching of cases and controls was addressed<br>*Cross-sectional study* - If applicable, describe analytical methods taking account of sampling strategy<br>(e) Describe any sensitivity analyses | p. 7-8 |  |  |
| Data access and cleaning methods |  | .. |  | RECORD 12.1: Authors should describe the extent to which the investigators had access to the | p. 6 |

**Supplemental Table 1.** RECORD Checklist
|  | Item No. | STROBE items | Location in manuscript where items are reported | RECORD items | Location in manuscript where items are reported |
| --- | --- | --- | --- | --- | --- |
|  |  |  |  | database population used to create the study population. |  |
|  |  |  |  | RECORD 12.2: Authors should provide information on the data cleaning methods used in the study. |  |
| Linkage |  | .. |  | RECORD 12.3: State whether the study included person-level, institutional-level, or other data linkage across two or more databases. The methods of linkage and methods of linkage quality evaluation should be provided. | p. 6 |
| **Results** |  |  |  |  |  |
| Participants | 13 | (a) Report the numbers of individuals at each stage of the study (*e.g.*, numbers potentially eligible, examined for eligibility, confirmed eligible, included in the study, completing follow-up, and analysed)<br>(b) Give reasons for non-participation at each stage.<br>(c) Consider use of a flow diagram | p. 8, 24 | RECORD 13.1: Describe in detail the selection of the persons included in the study (*i.e.*, study population selection) including filtering based on data quality, data availability and linkage. The selection of included persons can be described in the text and/or by means of the study flow diagram. | p. 8, 24 |
| Descriptive data | 14 | (a) Give characteristics of study participants (*e.g.*, | p. 8, 15-19 |  |  |

**Supplemental Table 1.** RECORD Checklist
|  | Item No. | STROBE items | Location in manuscript where items are reported | RECORD items | Location in manuscript where items are reported |
| --- | --- | --- | --- | --- | --- |
|  |  | demographic, clinical, social) and information on exposures and potential confounders<br>(b) Indicate the number of participants with missing data for each variable of interest<br>(c) *Cohort study* - summarise follow-up time (e.g., average and total amount) |  |  |  |
| Outcome data | 15 | *Cohort study* - Report numbers of outcome events or summary measures over time<br>*Case-control study* - Report numbers in each exposure category, or summary measures of exposure<br>*Cross-sectional study* - Report numbers of outcome events or summary measures | p. 8-9 |  |  |
| Main results | 16 | (a) Give unadjusted estimates and, if applicable, confounder-adjusted estimates and their precision (e.g., 95% confidence interval). Make | p. 8-9 |  |  |

**Supplemental Table 1.** RECORD Checklist
|  | Item No. | STROBE items | Location in manuscript where items are reported | RECORD items | Location in manuscript where items are reported |
| --- | --- | --- | --- | --- | --- |
|  |  | clear which confounders were adjusted for and why they were included<br>(b) Report category boundaries when continuous variables were categorized<br>(c) If relevant, consider translating estimates of relative risk into absolute risk for a meaningful time period |  |  |  |
| Other analyses | 17 | Report other analyses done—e.g., analyses of subgroups and interactions, and sensitivity analyses | p. 8-9 |  |  |
| **Discussion** |  |  |  |  |  |
| Key results | 18 | Summarise key results with reference to study objectives | p. 10 |  |  |
| Limitations | 19 | Discuss limitations of the study, taking into account sources of potential bias or imprecision. Discuss both direction and magnitude of any potential bias | p. 12 | RECORD 19.1: Discuss the implications of using data that were not created or collected to answer the specific research question(s). Include discussion of misclassification bias, unmeasured confounding, missing data, and changing eligibility over | p. 12 |

**Supplemental Table 1.** RECORD Checklist
|  | Item No. | STROBE items | Location in manuscript where items are reported | RECORD items | Location in manuscript where items are reported |
| --- | --- | --- | --- | --- | --- |
|  |  |  |  | time, as they pertain to the study being reported. |  |
| Interpretation | 20 | Give a cautious overall interpretation of results considering objectives, limitations, multiplicity of analyses, results from similar studies, and other relevant evidence | p. 10,11 |  |  |
| Generalisability | 21 | Discuss the generalisability (external validity) of the study results | p. 10,11 |  |  |
| **Other Information** |  |  |  |  |  |
| Funding | 22 | Give the source of funding and the role of the funders for the present study and, if applicable, for the original study on which the present article is based | p. 1 |  |  |
| Accessibility of protocol, raw data, and programming code |  | .. |  | RECORD 22.1: Authors should provide information on how to access any supplemental information such as the study protocol, raw data, or programming code. | p. 2 |

**Supplemental Table 2.**
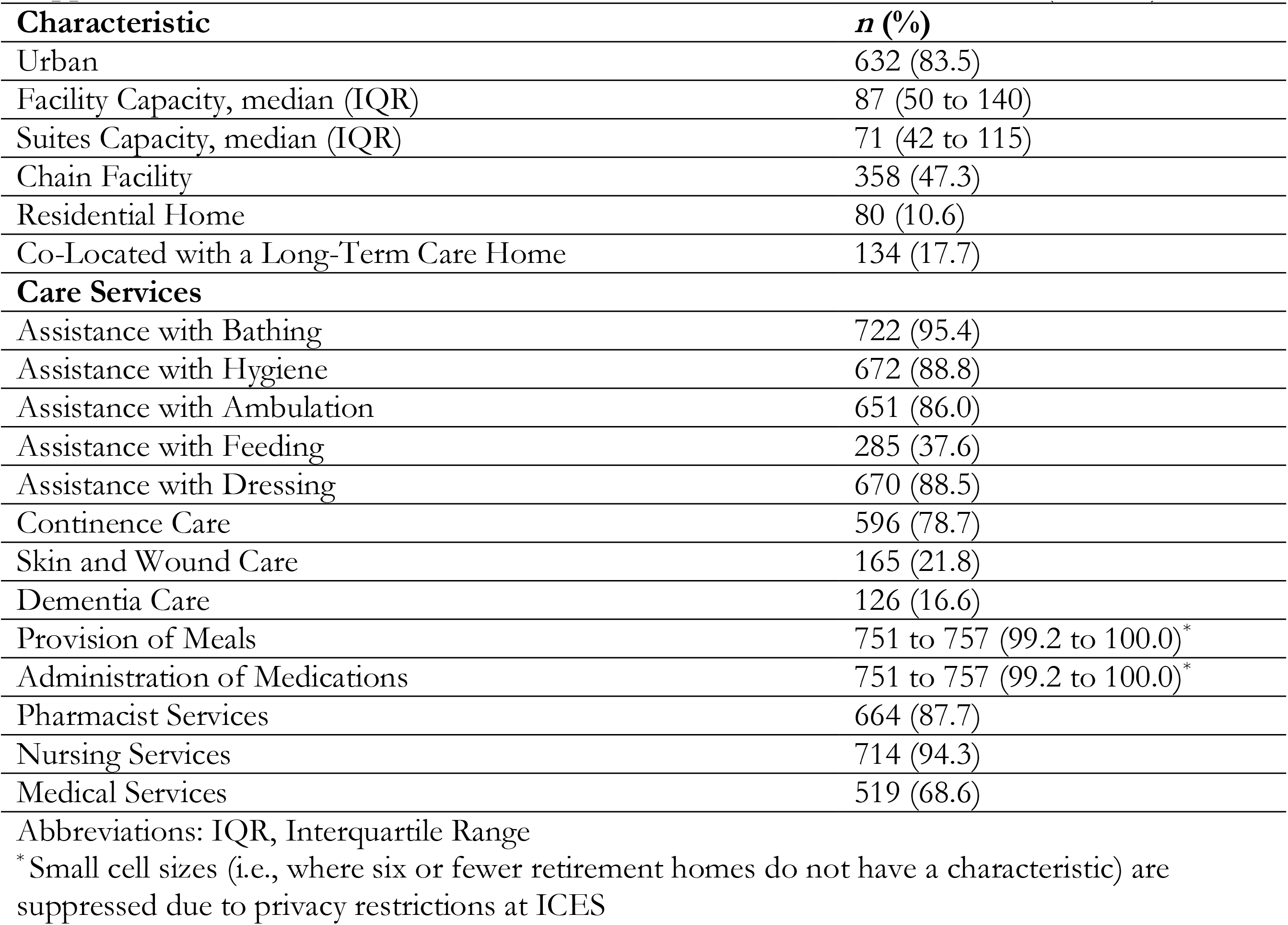
Characteristics of Licensed Retirement Homes in 2018 (*n* = 757)

**Supplemental Table 3.** Descriptions of the Health System Administrative Databases

| **Database** | **Description** |
| --- | --- |
| Client Agency Program Enrolment Data (CAPE) | Contains enrolment data on individuals in Ontario who are eligible for coverage under the universal health insurance plan in the province and enrolled or rostered to a health care program with a specific practitioner or group. |
| Continuing Care Reporting System (CCRS) | Contains person-level data (i.e., clinical, demographic) in nursing homes in Canada using the validated Resident Assessment Instrument Minimum Data Set (RAI-MDS) version 2.0. Complete assessments are conducted when people are admitted to the nursing home, every year thereafter, and after any significant change in the person's health by a health care provider. |
| Corporate Provider Database (CPDB) | Contains information on physician and allied health care practitioners (i.e., demographic characteristics, practice location, billing status and specialty) and/or groups (i.e., hospital groups and affiliated institutions, geographic location, group affiliation and composition) that are eligible to receive payments under the universal health insurance plan in Ontario. |
| Discharge Abstract Database (DAD) | Contains person-level data (i.e., demographic, administrative, diagnoses, procedures) for all admissions to acute care hospitals. The DAD is compiled and maintained by the Canadian Institute for Health Information. |
| Home Care Database (HCD) | Contains person-level data (i.e., social, clinical) for publicly funded home care services in Ontario, including types and volume of service provision. |
| National Ambulatory Care Reporting System (NACRS) | Contains person-level data (i.e., demographic, administrative, diagnoses, procedures) for all patient visits to ambulatory care centres in hospitals and communities (i.e., emergency departments, day surgery units, hemodialysis units, cancer care clinics). The NACRS is compiled and maintained by the Canadian Institute for Health Information. |
| Ontario Health Insurance Plan (OHIP) Database | Contains physician billings claims data among physicians who are remunerated by fee-for-service for outpatient visits. It also contains "shadow billings" for physicians who are remunerated through alternate payment schemes. |
| Ontario Marginalization Index (ONMARG) | Contains geographic, Census-based indices that quantify the extent of marginalization in communities in Ontario. There are four dimensions: residential instability, material deprivation, dependency, and ethnic concentration. |
| Postal Code Conversion File (PCCF+) | Specialized macro containing geographic identifiers based on Census data. This macro is based on 2016 Census information, identifies communities with less than 10,000 residents as rural, and includes related data from Canada Post Corporation. |
| Registered Persons Database (RPDP) | Contains demographic information (i.e., sex, age, date of birth, date of death for deceased individuals, area of residence, including postal code) and establishes eligibility for publicly funded universal health insurance in Ontario. |

**Supplemental Table 4.** Annual, Standardized Health Service Rates Among Residents of Retirement Homes, Residents of Long-Term Care Homes, and Home Care Recipients in the Community in 2018

|  |  | **ED Visits <sup>1</sup>** | **Hospitalizations <sup>1</sup>** | **ALC Days <sup>1</sup>** | **Primary Care Visits <sup>1</sup>** | **Specialist Physician Visits <sup>1</sup>** |
| --- | --- | --- | --- | --- | --- | --- |
| Total | Crude Rate | 0.0001 (0.0001 to 0.029231) | 0.0001 (0.0001 to 0.0001) | 0.0001 (0.0001 to 0.0001) | 0.0576923 (0.0192308 to 0.0961538) | 0.0192308 (0.001 to 0.576923) |
|  | Residents of Retirement Homes | 0.019337 (0.0001 to 0.07) | 0.0001 (0.0001 to 0.0216049) | 0.0001 (0.0001 to 0.0001) | 0.0769231 (0.0192308 to 0.1573034) | 0.0192308 (0.0001 to 0.0576923) |
|  | Residents of Long-Term Care Homes | 0.0001 (0.0001 to 0.0001) | 0.0001 (0.0001 to 0.0001) | 0.0001 (0.0001 to 0.0001) | 0.2307692 (0.2178988 to 0.2763158) | 0.0001 (0.0001 to 0.0001) |
|  | Home Care Recipients in the Community | 0.0001 (0.0001 to 0.0551181) | 0.0001 (0.0001 to 0.0001) | 0.0001 (0.0001 to 0.0001) | 0.0514706 (0.0001 to 0.1555556) | 0.0269231 (0.0001 to 0.1609195) |
|  | Community-Dwelling Older Adults | 0.0001 (0.0001 to 0.0001) | 0.0001 (0.0001 to 0.0001) | 0.0001 (0.0001 to 0.0001) | 0.0384615 (0.0192308 to 0.0961538) | 0.0192308 (0.0001 to 0.0384615) |
| Males | Crude Rate | 0.0001 (0.0001 to 0.0192308) | 0.0001 (0.0001 to 0.0001) | 0.0001 (0.0001 to 0.0001) | 0.0393258 (0.0192308 to 0.0961538) | 0.0192308 (0.0001 to 0.0576923) |
|  | Residents of Retirement Homes | 0.0233333 (0.0001 to 0.0823529) | 0.0001 (0.0001 to 0.0290456) | 0.0001 (0.0001 to 0.0001) | 0.0769231 (0.0192308 to 0.1655405) | 0.0192308 (0.0001 to 0.0769231) |
|  | Residents of Long-Term Care Homes | 0.0001 (0.0001 to 0.0191208) | 0.0001 (0.0001 to 0.0001) | 0.0001 (0.0001 to 0.0001) | 0.2307692 (0.2115385 to 0.2884615) | 0.0001 (0.0001 to 0.0192308) |
|  | Home Care Recipients in the Community | 0.0001 (0.0001 to 0.0707071) | 0.0001 (0.0001 to 0.0001) | 0.0001 (0.0001 to 0.0001) | 0.0492958 (0.0001 to 0.168) | 0.0384615 (0.0001 to 0.1891892) |
|  | Community-Dwelling Older Adults | 0.0001 (0.0001 to 0.0001) | 0.0001 (0.0001 to 0.0001) | 0.0001 (0.0001 to 0.0001) | 0.0384615 (0.0192308 to 0.0769231) | 0.0192308 (0.0001 to 0.0576923) |
| Females | Crude Rate | 0.0001 (0.0001 to 0.0192308) | 0.0001 (0.0001 to 0.0001) | 0.0001 (0.0001 to 0.0001) | 0.0576923 (0.0192308 to 0.1153846) | 0.0192308 (0.0001 to 0.0576923) |

|  | **ED Visits <sup>1</sup>** | **Hospitalizations <sup>1</sup>** | **ALC Days <sup>1</sup>** | **Primary Care Visits <sup>1</sup>** | **Specialist Physician Visits <sup>1</sup>** |
| --- | --- | --- | --- | --- | --- |
| Residents of Retirement Homes | 0.0192308 (0.0001 to 0.0644172) | 0.0001 (0.0001 to 0.0196629) | 0.0001 (0.0001 to 0.0001) | 0.0769231 (0.0192308 to 0.1538462) | 0.0001 (0.0001 to 0.0576923) |
| Residents of Long-Term Care Homes | 0.0001 (0.0001 to 0.0001) | 0.0001 (0.0001 to 0.0001) | 0.0001 (0.0001 to 0.0001) | 0.2307692 (0.2210526 to 0.2705314) | 0.0001 (0.0001 to 0.0001) |
| Home Care Recipients in the Community | 0.0001 (0.0001 to 0.0384615) | 0.0001 (0.0001 to 0.0001) | 0.0001 (0.0001 to 0.0001) | 0.0532319 (0.0001 to 0.15) | 0.0192308 (0.0001 to 0.14) |
| Community-Dwelling Older Adults | 0.0001 (0.0001 to 0.0001) | 0.0001 (0.0001 to 0.0001) | 0.0001 (0.0001 to 0.0001) | 0.0576923 (0.0192308 to 0.0961538) | 0.0192308 (0.0001 to 0.0384615) |

Abbreviations: ALC, Alternate Levels of Care; ED, Emergency Department
<sup>1</sup> Medians and Interquartile Ranges are presented

**Supplemental Table 5.** Adjusted Beta Coefficients (Standard Errors) from Annual, Standardized Health Service Rates Among Residents of Retirement Homes, Residents of Long-Term Care Homes, and Home Care Recipients in the Community in 2018

|  | ED Visits | Hospitalizations | ALC Days | Primary Care Visits | Specialist Physician Visits |
| --- | --- | --- | --- | --- | --- |
| **Demographic Characteristics** |  |  |  |  |  |
| Age | 0.002 (0.000) * | 0.001 (0.000) * | 0.034 (0.000) * | -0.002 (0.000) * | -0.018 (0.000) * |
| Sex |  |  |  |  |  |
| Male | (Ref) | (Ref) | (Ref) | (Ref) | (Ref) |
| Female | -0.026 (0.003) * | -0.163 (0.003) * | -0.092 (0.002) * | -0.023 (0.002) * | -0.100 (0.003) * |
| **Clinical Comorbidities** |  |  |  |  |  |
| Asthma | 0.159 (0.004) * | 0.089 (0.004) * | -0.115 (0.003) * | 0.160 (0.003) * | 0.131 (0.004) * |
| Cancer | 0.238 (0.004) * | 0.694 (0.003) * | 0.645 (0.003) * | 0.178 (0.003) * | 0.222 (0.003) * |
| Cardiac Arrhythmias | 0.301 (0.004) * | 0.335 (0.003) * | 0.138 (0.003) * | 0.119 (0.003) * | 0.052 (0.005) * |
| Cerebrovascular Accident | 0.251 (0.005) * | 0.525 (0.005) * | 1.080 (0.004) * | 0.005 (0.004) | 0.052 (0.005) * |
| Chronic Coronary Disease | 0.188 (0.003) * | 0.238 (0.003) * | -0.108 (0.003) * | 0.064 (0.002) * | 0.158 (0.003) * |
| Chronic Obstructive Pulmonary Disease | 0.267 (0.003) * | 0.329 (0.003) * | 0.256 (0.003) * | 0.079 (0.003) * | 0.054 (0.003) * |
| Congestive Heart Failure | 0.254 (0.005) * | 0.630 (0.004) * | 0.787 (0.004) * | 0.001 (0.004) | 0.072 (0.005) * |
| Dementia | -0.107 (0.006) * | 0.007 (0.005) | 1.083 (0.004) * | -0.138 (0.004) * | -0.237 (0.005) * |
| Diabetes | 0.060 (0.003) * | 0.079 (0.003) * | 0.124 (0.002) * | 0.215 (0.002) * | 0.075 (0.003) * |
| Hypertension | 0.134 (0.003) * | 0.238 (0.003) * | 0.088 (0.003) * | 0.298 (0.002) * | 0.132 (0.003) * |
| Mood Disorders | 0.203 (0.003) * | 0.023 (0.003) * | 0.126 (0.002) * | 0.222 (0.002) * | 0.192 (0.003) * |
| Myocardial Infarction | 0.207 (0.006) * | 0.473 (0.005) * | 0.302 (0.004) * | -0.030 (0.004) * | -0.005 (0.005) |
| Osteoarthritis | 0.155 (0.003) * | 0.326 (0.003) * | -0.140 (0.002) * | 0.212 (0.002) * | 0.287 (0.003) * |
| Osteoporosis | 0.003 (0.004) | -0.062 (0.004) * | 0.150 (0.003) * | 0.174 (0.003) * | 0.191 (0.004) * |
| Other Mental Health | 0.165 (0.003) * | 0.122 (0.003) * | 0.397 (0.003) * | 0.161 (0.002) * | 0.228 (0.003) * |
| Renal Disease | 0.223 (0.005) * | 0.440 (0.005) * | 0.665 (0.004) * | 0.056 (0.004) * | 0.475 (0.005) * |
| Rheumatoid Arthritis | 0.166 (0.008) * | 0.196 (0.008) * | 0.307 (0.007) * | 0.056 (0.006) * | 0.301 (0.008) * |
| **Community Characteristics** |  |  |  |  |  |
| Location |  |  |  |  |  |
| Rural | (Ref) | (Ref) | (Ref) | (Ref) | (Ref) |
| Urban | -0.532 (0.004) * | -0.126 (0.004) * | -0.152 (0.003) * | 0.289 (0.003) * | 0.243 (0.004) * |
| Neighborhood Income Quintile |  |  |  |  |  |

|  | **ED Visits** | **Hospitalizations** | **ALC Days** | **Primary Care Visits** | **Specialist Physician Visits** |
| --- | --- | --- | --- | --- | --- |
| 1 (Least) | (Ref) | (Ref) | (Ref) | (Ref) | (Ref) |
| 2 | -0.084 (0.004) * | -0.040 (0.004) * | -0.289 (0.003) * | 0.032 (0.003) * | 0.081 (0.004) * |
| 3 | -0.121 (0.005) * | -0.069 (0.004) * | -0.422 (0.004) * | 0.050 (0.003) * | 0.124 (0.004) * |
| 4 | -0.168 (0.005) * | -0.071 (0.005) * | -0.419 (0.004) * | 0.062 (0.004) * | 0.167 (0.005) * |
| 5 (Highest) | -0.226 (0.005) * | -0.065 (0.005) * | -0.447 (0.004) * | 0.053 (0.004) * | 0.257 (0.005) * |
| Ontario Marginalization Index Summary Score | -0.043 (0.002) * | -0.042 (0.002) * | -0.009 (0.002) * | 0.033 (0.002) * | 0.062 (0.002) * |
| Constant | -5.016 (0.017) * | -7.243 (0.016) * | -10.781 (0.013) * | -3.694 (0.013) * | -3.364 (0.016) * |

Abbreviations: ALC, Alternate Levels of Care; ED, Emergency Department
\* $P < .001$

**Supplemental Table 6.**
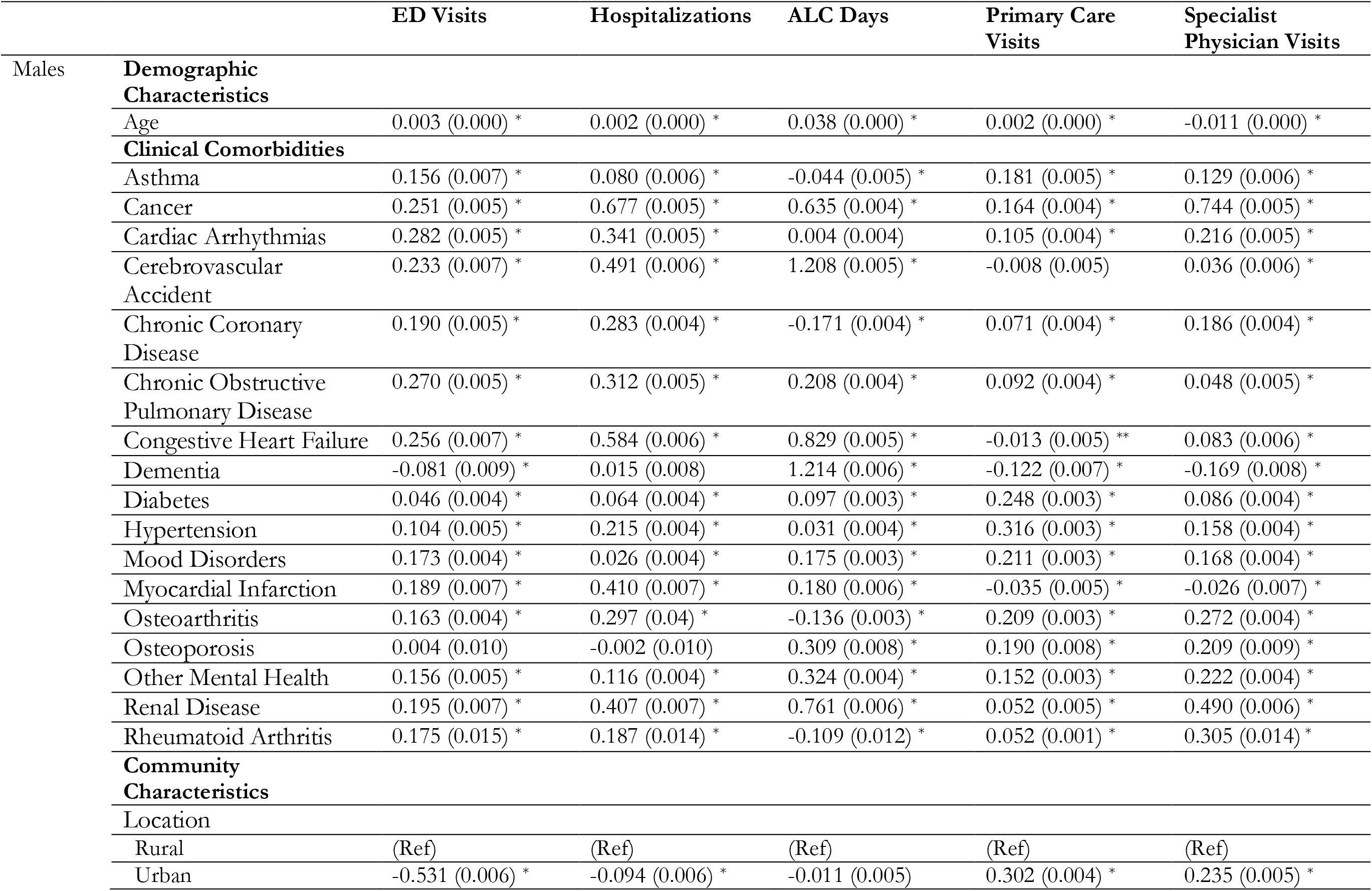
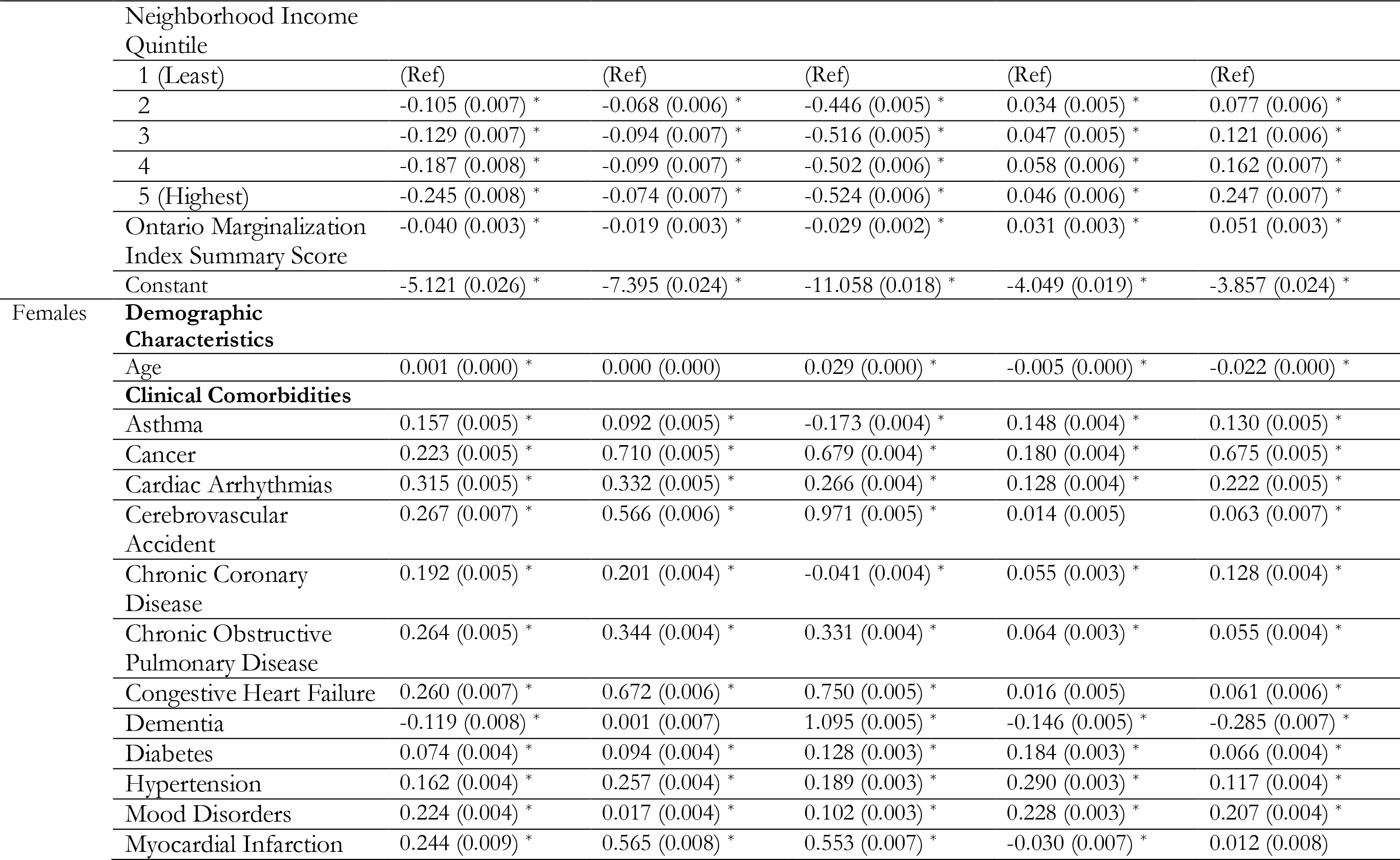
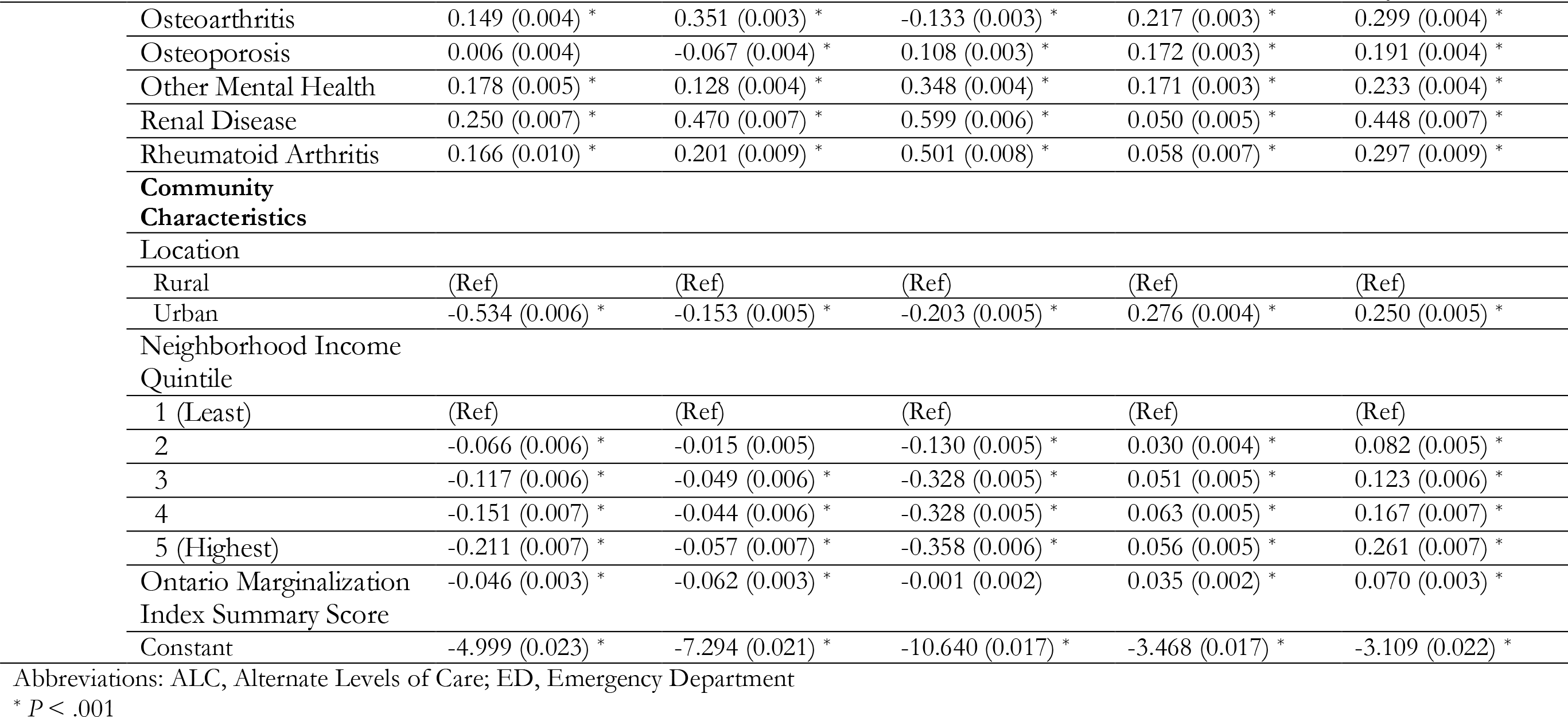
Sex-Stratified Subgroup Analysis Adjusted Beta Coefficients (Standard Errors) from Annual, Standardized Health Service Rates Among Residents of Retirement Homes, Residents of Long-Term Care Homes, and Home Care Recipients in the Community in 2018

## REFERENCES

1. Poss JW, Sinn C-LJ, Grinchenko G, Blums J, Peirce T, Hirdes J. Location, Location, Location: Characteristics and Services of Long-Stay Home Care Recipients in Retirement Homes Compared to Others in Private Homes and Long-Term Care Homes. Healthc Policy Polit Sante. 2017;12(3):80–93.

2. Roblin B, Deber R, Kuluski K, Silver MP. Ontario’s Retirement Homes and Long-Term Care Homes: A Comparison of Care Services and Funding Regimes. Can J Aging. 2019 Jun;38(2):155–67.

3. Temkin-Greener H, Mao Y, Ladwig S, Cai X, Zimmerman S, Li Y. Variability and Potential Determinants of Assisted Living State Regulatory Stringency. J Am Med Dir Assoc. 2021;22(8):1714-1719.e2.

4. Poss J, Sinn C-L, Grinchenko G, Salam-White L, Hirdes J. Comparing Changes and Transitions of Home Care Clients in Retirement Homes and Private Homes. Can J Aging. 2019 Aug 15;1–11.

5. Simmons SF, Schnelle JF, Sathe NA, Slagle JM, Stevenson DG, Carlo ME, et al. Defining Safety in the Nursing Home Setting: Implications for Future Research. J Am Med Dir Assoc. 2016 Jun 1;17(6):473–81.

6. Cornell PY, Zhang W, Thomas KS. Changes in Long-Term Care Markets: Assisted Living Supply and the Prevalence of Low-Care Residents in Nursing Homes. J Am Med Dir Assoc. 2020 Aug;21(8):1161-1165.e4.

7. Grabowski DC, Stevenson DG, Cornell PY. Assisted living expansion and the market for nursing home care. Health Serv Res. 2012 Dec;47(6):2296–315.

8. Han K, Trinkoff AM, Storr CL, Lerner N, Yang BK. Variation Across U.S. Assisted Living Facilities: Admissions, Resident Care Needs, and Staffing. J Nurs Scholarsh Off Publ Sigma Theta Tau Int Honor Soc Nurs. 2017;49(1):24–32.

9. Thomas KS, Zhang W, Cornell PY, Smith L, Kaskie B, Carder PC. State Variability in the Prevalence and Healthcare Utilization of Assisted Living Residents with Dementia. J Am Geriatr Soc. 2020;68(7):1504–11.

10. Arneson L, Bender AA, Robert MN, Perkins MM. Optimizing Quality of Life With Cognitive Impairment: A Study of End-of-Life Care in Assisted Living. J Am Med Dir Assoc. 2020 May;21(5):692–6.

11. Bartley MM, Quigg SM, Chandra A, Takahashi PY. Health Outcomes From Assisted Living Facilities: A Cohort Study of a Primary Care Practice. J Am Med Dir Assoc. 2018 Mar;19(3).

12. Thomas KS, Dosa D, Gozalo PL, Grabowski DC, Nazareno J, Makineni R, et al. A Methodology to Identify a Cohort of Medicare Beneficiaries Residing in Large Assisted Living Facilities Using Administrative Data. Med Care. 2018;56(2):e10–5.

13. Maxwell CJ, Soo A, Hogan DB, Wodchis WP, Gilbart E, Amuah J, et al. Predictors of Nursing Home Placement from Assisted Living Settings in Canada. Can J Aging Rev Can Vieil. 2013 Dec;32(4):333–48.

14. Maxwell CJ, Amuah JE, Hogan DB, Cepoiu-Martin M, Gruneir A, Patten SB, et al. Elevated Hospitalization Risk of Assisted Living Residents With Dementia in Alberta, Canada. J Am Med Dir Assoc. 2015;16(7):568–77.

15. Rockwood JK, Richard M, Garden K, Hominick K, Mitnitski A, Rockwood K. Precipitating and predisposing events and symptoms for admission to assisted living or nursing home care. Can Geriatr J CGJ. 2014;17(1):16–21.

16. Manis DR, Rahim A, Poss JW, Bielska IA, Bronskill SE, Tarride J-E, et al. Association Between Dementia Care Programs in Assisted Living Facilities and Transitions to Nursing Homes in Ontario, Canada: A Population-Based Cohort Study. J Am Med Dir Assoc. 2021;(100893243).

17. Manis DR, Rahim A, Poss JW, Bielska IA, Bronskill SE, Tarride J-É, et al. Do assisted living facilities that offer a dementia care program differ from those that do not? A population-level cross-sectional study in Ontario, Canada. BMC Geriatr. 2021 Aug 16;21(1):463.

18. Benchimol EI, Smeeth L, Guttmann A, Harron K, Moher D, Petersen I, et al. The REporting of studies Conducted using Observational Routinely-collected health Data (RECORD) Statement. PLOS Med. 2015 Oct 6;12(10):e1001885.

19. Brath H, Kim SJ, Bronskill SE, Rochon PA, Stall NM. Co-locating Older Retirement Home Residents: Uncovering an Under-Researched Population via Postal Code. Healthc Policy. 2020 Nov 10;16(2):69–81.

20. Austin PC, Daly PA, Tu JV. A multicenter study of the coding accuracy of hospital discharge administrative data for patients admitted to cardiac care units in Ontario. Am Heart J. 2002;144(2):290–6.

21. Gershon AS, Wang C, Guan J, Vasilevska-Ristovska J, Cicutto L, To T. Identifying Individuals with Physcian Diagnosed COPD in Health Administrative Databases. COPD J Chronic Obstr Pulm Dis. 2009;6(5):388–94.

22. Gershon AS, Wang C, Guan J, Vasilevska-Ristovska J, Cicutto L, To T. Identifying patients with physician-diagnosed asthma in health administrative databases. Can Respir J J Can Thorac Soc. 2009;16(6):183–8.

23. Hux JE, Ivis F, Flintoft V, Bica A. Diabetes in Ontario: Determination of prevalence and incidence using a validated administrative data algorithm. Diabetes Care. 2002 Mar 1;25(3):512–6.

24. Jaakkimainen RL, Bronskill SE, Tierney MC, Herrmann N, Green D, Young J, et al. Identification of Physician-Diagnosed Alzheimer’s Disease and Related Dementias in Population-Based Administrative Data: A Validation Study Using Family Physicians’ Electronic Medical Records. J Alzheimers Dis. 2016 Jan 1;54(1):337–49.

25. Schultz SE, Rothwell DM, Chen Z, Tu K. Identifying cases of congestive heart failure from administrative data: a validation study using primary care patient records. Chronic Dis Inj Can. 2013 Jun;33(3):160–6.

26. Tu K, Campbell NR, Chen Z-L, Cauch-Dudek KJ, McAlister FA. Accuracy of administrative databases in identifying patients with hypertension. Open Med. 2007 Apr 14;1(1):e18–26.

27. Widdifield J, Bernatsky S, Paterson JM, Tu K, Ng R, Thorne JC, et al. Accuracy of Canadian Health Administrative Databases in Identifying Patients With Rheumatoid Arthritis: A Validation Study Using the Medical Records of Rheumatologists. Arthritis Care Res. 2013;65(10):1582–91.

28. Hardin JW, Hilbe JM. Generalized Linear Models and Extensions. 4th ed. College Station, Texas, USA: Stata Press; 2018.

29. Allin S, Rudoler D, Mullen J. Experiences with Two-Tier Home Care in Canada: A Focus on Inequalities in Home Care Use by Income in Ontario. In: Flood CM, Thomas B, editors. Is Two-Tier Health Care the Future? Ottawa: University of Ottawa Press; 2020. p. 123–44.

30. Hurley J. Borders, Fences, and Crossings: Regulating Parallel Private Finance in Health Care. In: Flood CM, Thomas B, editors. Is Two-Tier Health Care the Future? Ottawa: University of Ottawa Press; 2020. p. 69–91.

31. Hua CL, Zhang W, Cornell PY, Rahman M, Dosa DM, Thomas KS. Characterizing Emergency Department Use in Assisted Living. J Am Med Dir Assoc. 2021 Apr 1;22(4):913-917.e2.

32. Carder PC. State Regulatory Approaches for Dementia Care in Residential Care and Assisted Living. The Gerontologist. 2017;57(4):776–86.

33. McGregor MJ, McGrail KM, Abu-Laban RB, Ronald LA, Baumbusch J, Andrusiek D, et al. Emergency department visit rates and patterns in Canada’s Vancouver coastal health region. Can J Aging Rev Can Vieil. 2014 Jun;33(2):154–62.

34. Canadian Mortgage and Housing Corporation. Results from the 2020 Seniors Housing Survey [Internet]. 2020 [cited 2021 Jul 15]. Available from: https://www.cmhc-schl.gc.ca/en/blog/2020-housing-observer/results-2020-seniors-housing-survey

35. Pearson CF, Quinn CC, Loganathan S, Datta AR, Mace BB, Grabowski DC. The Forgotten Middle: Many Middle-Income Seniors Will Have Insufficient Resources For Housing And Health Care. Health Aff Proj Hope. 2019;38(5):101377hlthaff201805233.

36. Stevenson DG, Grabowski DC. Sizing up the market for assisted living. Health Aff (Millwood). 2010 Feb;29(1):35–43.

37. Bowblis JR. Nursing home prices and market structure: the effect of assisted living industry expansion. Health Econ Policy Law. 2014;9(1):95–112.

38. Silver BC, Grabowski DC, Gozalo PL, Dosa D, Thomas KS. Increasing Prevalence of Assisted Living as a Substitute for Private-Pay Long-Term Nursing Care. Health Serv Res. 2018;53(6):4906–20

